# Who Gets Long COVID and Suffers its Mental Health and Socioeconomic Consequences in the United States? Preliminary Findings from a Large Nationwide Study

**DOI:** 10.1101/2023.01.06.23284199

**Authors:** Daniel Kim

## Abstract

As the number of confirmed COVID-19 cases now exceeds 100 million cases in the United States and continues to climb, concerns have been increasingly raised over the future public health and economic burden of long COVID including disability and concomitant declines in labor force participation. Only a handful of US population-based studies have explored sociodemographic and socioeconomic characteristics that put people at risk of long COVID or have investigated its mental health and socioeconomic sequelae. Herein, I report findings from the largest multivariable analysis to date using US nationally-representative data on 153,543 adults including 19,985 adults with long COVID to explore key predictors and sequelae of long COVID. An estimated 14.0% of adults aged 18-84 y (35.11 million adults) and 15.5% of working-aged adults aged 18-64 y (30.65 million adults) had developed long COVID by November 2022. Several sociodemographic and socioeconomic factors predicted long COVID including lower household income, being aged 30-49 y, Hispanic, female, gay/lesbian or bisexual, and divorced/separated. Even after accounting for such factors, having long COVID was linked to higher risks of recent unemployment, financial hardship, and anxiety and depressive symptomatology, with evidence of dose-response relationships. Overall, an estimated 27.7 million US adults aged 18-84 y and 24.2 million working-aged adults with long COVID who had been or may still be at risk of adverse socioeconomic and mental health outcomes. Lost work was further calculated to be the equivalent of 3 million workers annually, and the estimated annual lost earnings due to long COVID among working-aged adults totaled $175 billion. These preliminary findings highlight the substantial public health and economic implications of long COVID among Americans and should prompt further inquiry and intervention.

## INTRODUCTION

As the COVID-19 pandemic approaches the three-year mark and the number of confirmed cases now exceeds 100 million in the United States and continues to climb, concerns have been increasingly raised over the future public health and economic burden of long COVID including disability and concomitant declines in labor force participation. Long COVID (also known as post-COVID-19 syndrome) is a condition characterized by long-term health problems persisting beyond the typical recovery period for COVID-19 (2). By yielding symptoms affecting daily functioning, long COVID can plausibly affect one’s ability to work, leading to loss of employment income and inducing financial strain. Through impacting physical and mental functioning (3), long COVID can contribute to mental health disorders including anxiety and depression (4). Yet only a handful of US population-based studies have explored sociodemographic and socioeconomic predictors of long COVID and economic and health sequelae, and such studies remain limited to relatively small samples (5-10).

The present study was undertaken to use nationally-representative data from the US Census Bureau’s Household Pulse Survey (HPS) on 153,543 adults to investigate key sociodemographic and socioeconomic characteristics as risk factors for long COVID among adults, to explore the associations of long COVID with unemployment, financial hardship, and mental health outcomes, and to estimate the economic burden of lost wages from long COVID in the United States.

Repeated cross-sectional data were pooled from nationally-representative HPS surveys in data collection Phase 3.6 administered in 3 waves from September 14—November 14, 2022 (11). The HPS used the Census Bureau’s Master Address File as the source of sampled housing units. The HPS was conducted online by Qualtrics, with survey response rates ranging from 3.9-5.6% (12). The study population consisted of US adults aged 18–84 y.

Analyses of employment and financial hardship as outcomes were limited to adult participants of working age (aged 18-64 y). For samples analyzed in multivariable models, data were available on 153,543 adults including 113,192 working-aged adults (representative of 197 million American adults).

## MATERIALS AND METHODS

### Outcomes

The first set of multivariable models examined the risks of 1) long COVID in the full sample; 2) long COVID among those with COVID-19 infection; and 3) COVID-19 infection in the full sample. The risk of long COVID is a function of the risk of becoming infected with COVID and the risk of progressing from acute infection to long COVID. Long COVID was defined in the HPS as reporting any symptoms lasting 3 months or longer that were not present prior to having COVID-19 including fatigue, difficulty concentrating and forgetfulness, “brain fog”, shortness of breath, joint or muscle pain, heart palpitations, changes to taste/smell, and inability to exercise (12).

The second set of multivariable models examined 4 recent outcomes in the full sample and among those still reporting long COVID symptoms: whether the respondent was employed in the past week; experienced financial hardship, defined as household difficulty (somewhat/very difficult vs. not at all/a little difficult) to pay for usual household expenses including food, rent/ mortgage, and loans, within the previous week; and the frequency of experiencing anxiety symptoms and depressive symptoms (using the 2-item Generalized Anxiety Disorder-2 (GAD-2) and Patient Health Questionnaire-2 (PHQ-2)) over the previous 2 weeks, with scores of 3–6 vs 0–2 being employed previously to screen for anxiety and depressive disorders (10). Based on a cut-off score of ≥3, the GAD-2 demonstrated optimal sensitivity and specificity (sensitivity = 0.71, specificity = 0.69), and the PHQ-2 exhibited peak sensitivity and adequate specificity (sensitivity = 0.64, specificity = 0.85). Internal consistency reliability has been demonstrated to be high for both measures (Cronbach’s α = 0.81 and 0.83 for the GAD-2 and PHQ-2 scales, respectively) (13).

### Predictors

The first set of models included the following predictors: age group, gender (male, female, transgender, other), sexual orientation (straight, gay/lesbian, bisexual, other), race and Hispanic ethnicity (Hispanic, non-Hispanic White, Black, Asian, and Other), marital status, education, household income in 2021, household size, presence of children in the household, receipt of a COVID-19 primary vaccine or booster, and health insurance status (12).

In the second set of models, the main predictors were long COVID status (yes, no) in the full sample and the extent to which current long COVID symptoms reduced the ability to carry out day-to-day activities (a lot, a little, not at all) compared to before having COVID-19.

All models further controlled for self-reported non-receipt of the COVID-19 booster due to a COVID-19 infection and state of residence.

### Statistical analysis

Multivariable modified Poisson regression models (that can approximate yet circumvent issues with log-binomial models) were fit to estimate adjusted PRs from generalized estimating equations that accounted for repeated measures within individuals and survey weights and provided robust standard errors based on sandwich estimators (14). Multiple imputation using 25 multiply imputed datasets and the MCMC algorithm without rounding under a missing at random assumption (15) was used to handle missing data.

### Estimation of economic burden of lost wages

The estimated economic burden of long COVID from lost wages was calculated by adopting a similar approach to others (16). A typical 75% labor force participation rate was applied to the HPS-derived estimated number of adults aged 18-64 y with long COVID who still reported having long COVID symptoms. In these adults, I assumed that work time was reduced by 25% due to long COVID (16), and combined their estimated FTEs of lost work due to reduced working time with the lost FTEs for the HPS-derived number of adults choosing not to work recently because of long COVID. The total FTE of lost work was then multiplied by the average US wage of $1,106 per week to yield an annual estimate of lost earnings due to long COVID (16).

All analyses were conducted using SAS (version 9.4; SAS Institute, Cary, NC). All data were publicly available and anonymized and deemed exempt from ethical compliance.

## RESULTS

### Risk of long COVID in the full sample (n=153,543)

Compared to those aged 18-29 y, adults aged 30-39 y and 40-49 y had a 17-21% higher risk of long COVID, while adults aged 65+ y had a 31-48% lower risk (*Fig. 1*). Higher risks were also observed among those with any (vs. no) children in the household, of Hispanic and other (vs. White non-Hispanic) race/ethnicity, of lower income, in larger households, and who were divorced/separated (vs. married), gay/lesbian or bisexual (vs. straight), and female (52% higher risk), transgender (42% higher risk), or other gender (vs. male) (all multivariable-adjusted prevalence ratios, PRs = 1.08-1.77; nearly all *P*<0.05). Black and Asian adults and never married adults were at 21-33% lower risk and 8% lower risk than White adults and married adults, respectively (all *P*<0.05). No vaccinated groups exhibited lower risks than the unvaccinated group (all *P*>0.05) (*Fig. 1*).

**Figure 1.**
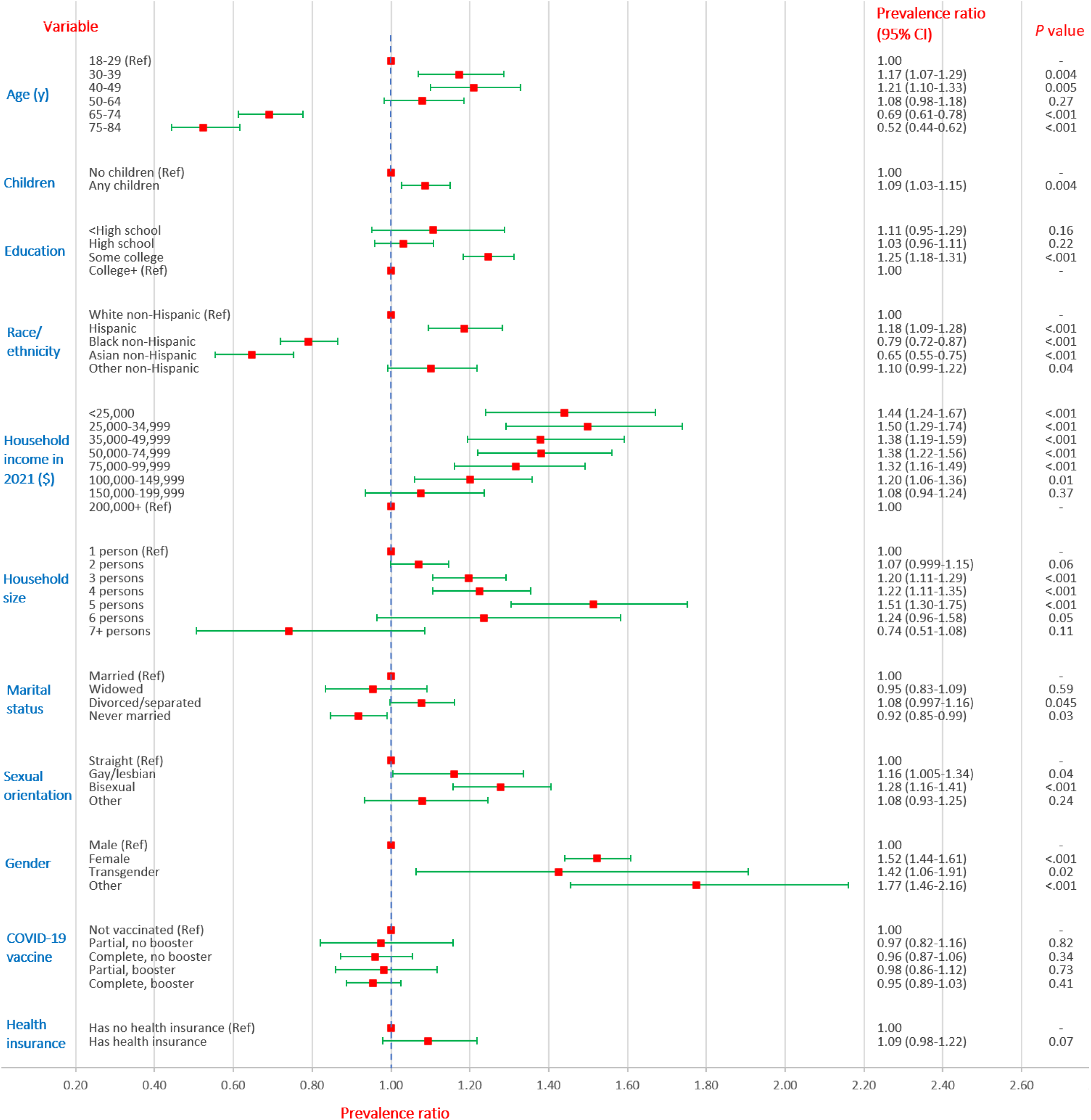
Multivariable-adjusted risks of long COVID according to sociodemographic and socioeconomic characteristics among 153,543 adults aged 18-84 y, U.S. Census Bureau Household Pulse Survey, September–November 2022. *All models included all listed predictor variables and were also adjusted for state of residence and non-receipt of a COVID-19 vaccine booster due to COVID-19 infection. ‘Partial’ refers to receipt of one dose of a two-shot primary vaccine series. ‘Complete’ refers to completion of the primary vaccine series (2 doses of a two-shot series or a single dose series).

### Risk of long COVID among those with COVID-19 infection (n=72,**636)**

Modeling the risk of long COVID among those with COVID-19 infection showed similar patterns (e.g., with females having a 37% higher risk than males; *Fig. 2*), with a few exceptions: Having a child in the household was not associated with long COVID, whereas inverse associations between educational attainment and long COVID were consistently observed. Being widowed was linked to a 14% higher risk, and no association was observed for never being married. Those who completed their primary vaccination series but did not receive a booster had a 15% higher risk than the unvaccinated (*Fig. 2*).

**Figure 2.**
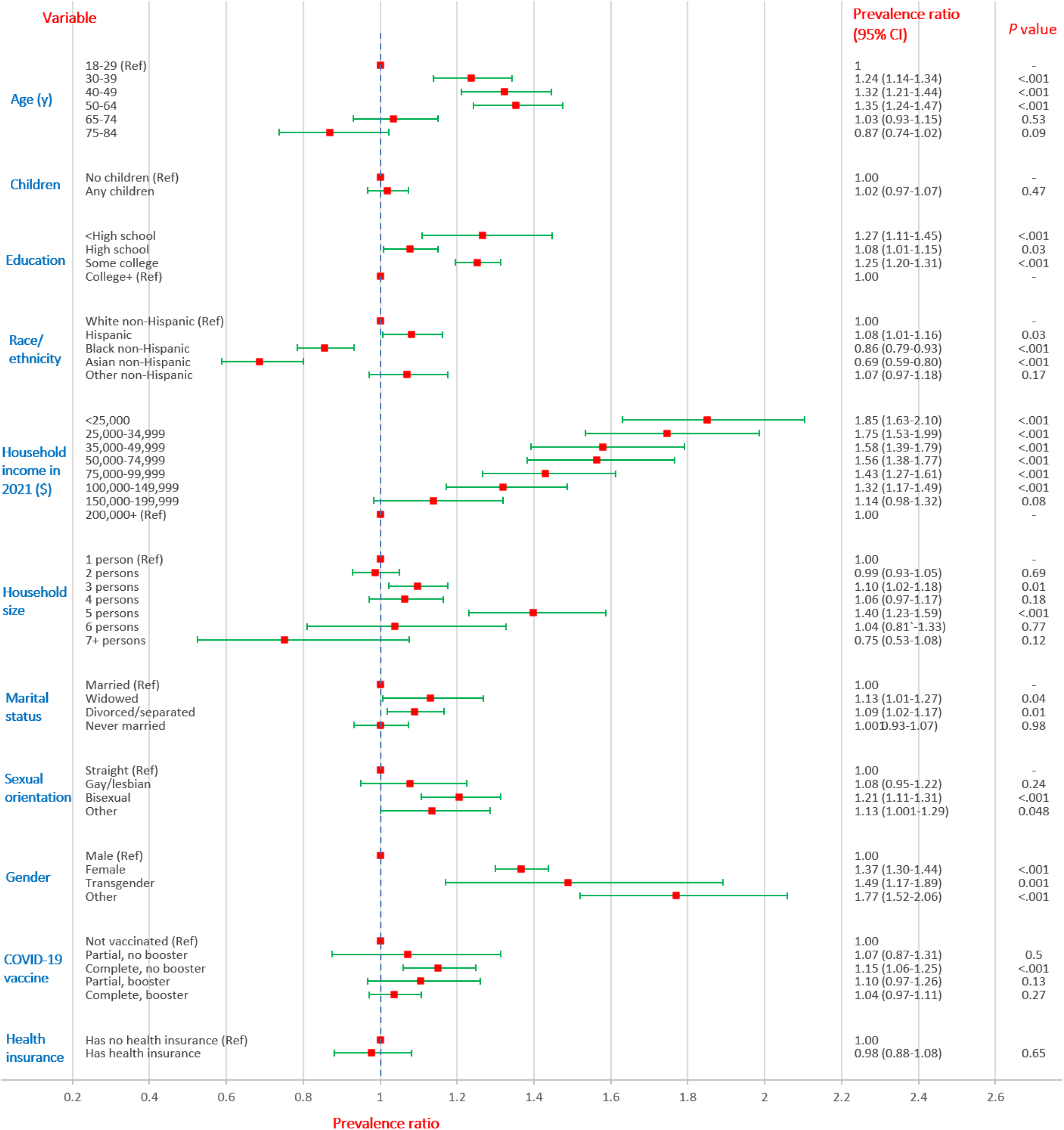
Multivariable-adjusted* risks of long COVID according to sociodemographic and socioeconomic characteristics among 72,636 adults aged 18-84 y with COVID-19 infection, U.S. Census Bureau Household Pulse Survey, September–November 2022. *All models included all listed predictor variables and were also adjusted for state of residence and non-receipt of a COVID-19 vaccine booster due to COVID-19 infection. ‘Partial’ refers to receipt of one dose of a two-shot primary vaccine series. ‘Complete’ refers to completion of the primary vaccine series (2 doses of a two-shot series or a single dose series

### Risk of COVID-19 infection in the full sample (n=153,**543)**

Modeling the risk of COVID-19 infection exhibited analogous patterns to the risk of long COVID (*Fig. 3*). However, being older more consistently predicted a lower risk of COVID-19, and lower education and income were associated with lower rather than higher risks. For gender, only females showed a higher risk of infection than males. All vaccinated groups had 9-17% lower risks than unvaccinated adults (all *P*<0.05) (*Fig. 3*).

**Figure 3.**
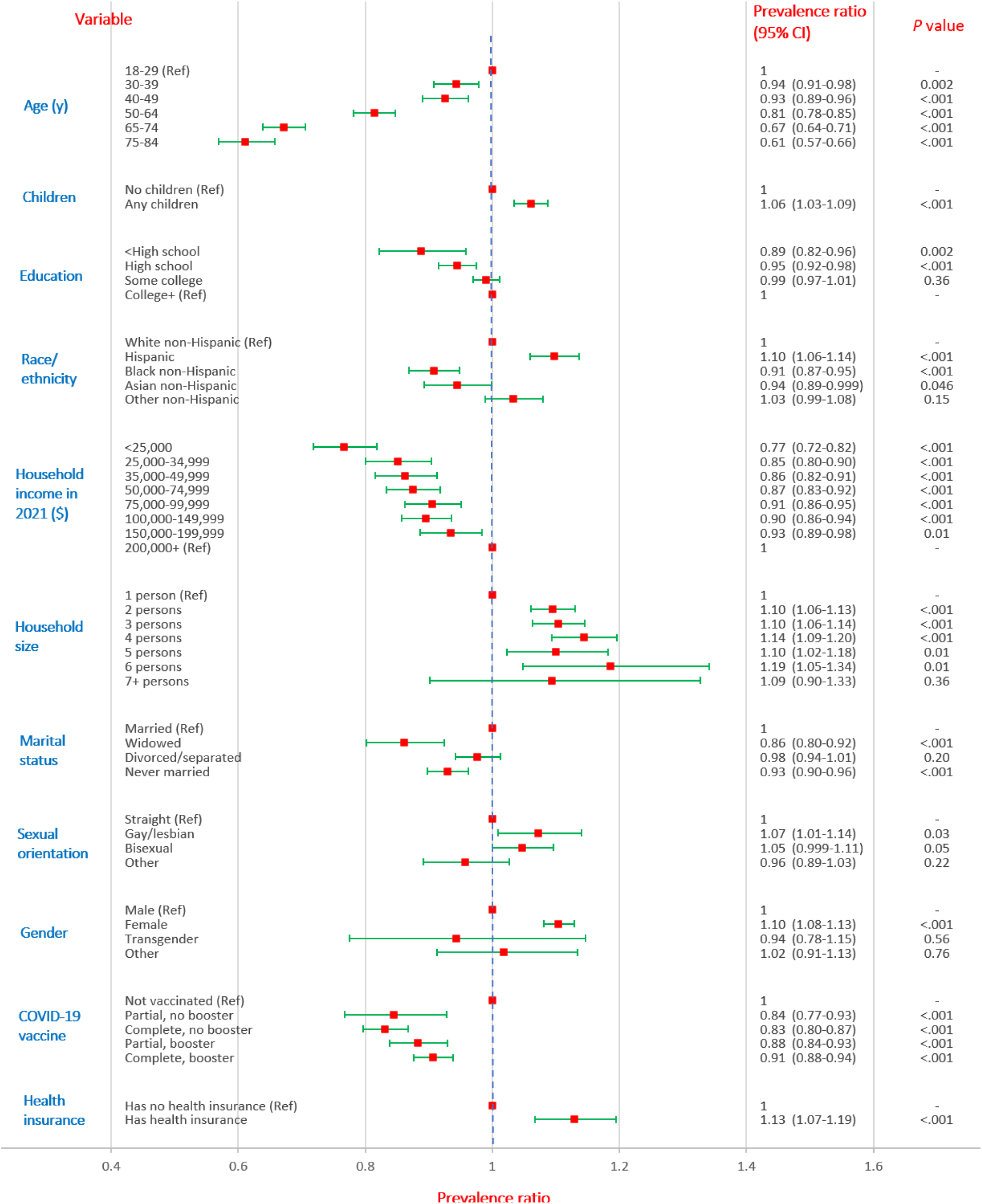
Multivariable-adjusted* risks of COVID-19 infection according to sociodemographic and socioeconomic characteristics among 153,543 adults aged 18-84 y, U.S. Census Bureau Household Pulse Survey, September–November 2022. *All models included all listed predictor variables and were also adjusted for state of residence and non-receipt of a COVID-19 vaccine booster due to COVID-19 infection. ‘Partial’ refers to receipt of one dose of a two-shot primary vaccine series. ‘Complete’ refers to completion of the primary vaccine series (2 doses of a two-shot series or a single dose series).

### Risk of unemployment, financial hardship, and anxiety and depression among working-aged adults (n=113,192) and among adults with current long COVID symptoms (n=10,348)

Those who reported having long COVID had a higher likelihood of not being employed, experiencing financial hardship, and reporting recent anxiety or depressive symptoms (PRs ranging from 1.05-1.51; all *P*<0.05 except for current employment; *Fig. 4*). Among those with current symptoms, there was evidence of dose-response relationships, with those reporting the highest (vs. no) impact on daily functioning having a more than two-fold higher risk of recent depressive symptoms (PR=2.23; 95%=1.92-2.60, p<0.001; *Fig. 4*).

**Figure 4.**
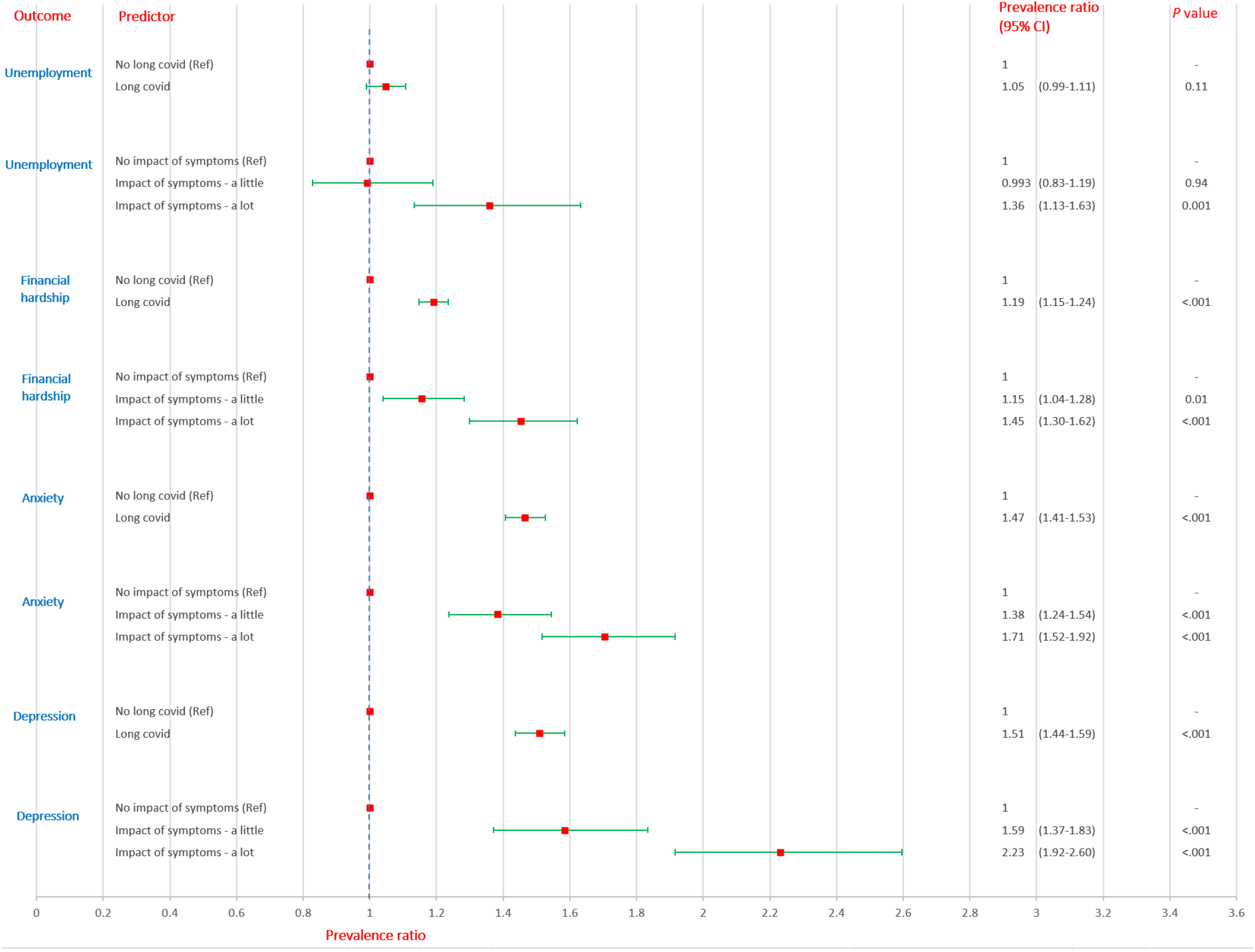
Multivariable-adjusted* risks of unemployment, financial hardship, and anxiety and depressive symptomatology according to long COVID status among 153,543 adults and according to impact of long COVID symptoms on daily functioning among 10,348 adults during the COVID-19 pandemic, U.S. Census Bureau Household Pulse Survey, September–November 2022. *All models were also adjusted for age, gender, race and Hispanic ethnicity, education, household income in 2021, sexual orientation, children in the household, household size, receipt of a COVID-19 vaccine, health insurance coverage, state of residence, and non-receipt of a vaccine booster due to a COVID-19 infection. The analyses of unemployment and financial hardship were restricted to 113,192 working-aged adults (aged 18-64 years). The analyses of anxiety and depression include all adults in the sample (aged 18-84 years). The impact of long COVID symptoms variable measured the extent to which current symptoms reduced the ability to carry out day-to-day activities compared with the period before having COVID-19.

### Estimated population at risk and economic burden of lost wages

Based on the study sample, 14.0% of US adults aged 18-84 y (35.11 million; 95% CI=34.18-36.04 million) and 15.5% of working-aged adults aged 18-64 y (30.65 million; 95% CI=29.77-31.53 million) had developed long COVID by November 2022. The estimated percentages of those with long COVID still experiencing symptoms and reporting a modest to considerable impact on daily functioning were 55.3% and 24.6%, respectively. Assuming the same percentages in those no longer experiencing symptoms, this translates into an estimated 28.1 million adults (95% CI=27.34-28.83 million) aged 18-84 y and 24.5 million working-aged adults (95% CI=23.81-25.22 million) with long COVID who had been or may still be at risk of adverse outcomes. Given that cases of COVID will continue to accrue from new infections and re-infections in the ongoing pandemic, these numbers of susceptible individuals will only escalate over time.

An estimated 14.5 million adults aged 18-64 y with long COVID still had current symptoms. Assuming a 75% labor force participation rate, while accounting for 435,473 American adults that reported not working recently because of long COVID, 10.4 million adults were working with long COVID. The total full-time equivalents (FTEs) of lost work either due to not working at all or 25% reduced working hours because of long COVID amounted to 3.05 million FTE. Using the average US wage per week, this translates into $175 billion annually in lost earnings due to long COVID for American adults aged 18-64 y—approximating a previous $168 billion estimate for adults aged 16-64 y (16). Notably, this figure likely substantially underestimates the total economic burden of long COVID as it does not account for the costs of lost productivity during work hours, reductions in quality of life, or the health care costs associated with long COVID and its physical and mental health sequelae which could be in excess of $500 billion annually (16,17).

## DISCUSSION

To the author’s knowledge, this study represents the largest multivariable analysis using nationally-representative data to explore key sociodemographic and socioeconomic predictors and mental health and economic sequelae and to estimate the economic burden of lost wages from long COVID in the United States. The large sample size enabled a comprehensive examination of potential risk factors including marital status and gender and the discovery of novel associations such as for sexual orientation. Several characteristics predicted greater long COVID risks including being aged 30-49 y, of lower education and household income, living in a larger household, and being Hispanic, female, gay/lesbian or bisexual, and divorced/separated. Receipt of a COVID-19 vaccine (with or without a booster) was linked to lower risks of infection.

Even after accounting for sociodemographic and socioeconomic factors, having long COVID predicted higher risks of recent unemployment, financial hardship, and anxiety and depressive symptomatology. For all of these outcomes, there were stronger relationships at higher levels of impacts of symptoms on daily functioning, compatible with causal relationships. Previous US population-based studies have likewise found long COVID to predict a lower odds of employment (9) and higher risk of anxiety (10).

Limitations of this study include its cross-sectional design, which precludes the ability to draw causal inferences due to bias from reverse causation or residual confounding by factors including comorbid conditions and reinfections (18). While adjustment was made for receipt of a vaccine booster due to a COVID-19 infection, confounding by indication could explain the null and positive associations between vaccination and long COVID. Finally, while sampling weights accounted for non-response and there is evidence that weighting adjustments mitigated non-response bias (19), such bias could have led to bias towards or away from the null.

The preliminary findings from this study highlight the anticipated substantial health and economic burden of long COVID among American adults. Should these estimates be confirmed in longitudinal studies, future efforts should focus on identifying biological and social explanations and mediating pathways, and on targeting risk stratification and risk reduction in those at higher risk of long COVID. In addition to ensuring dedicated funding for long COVID research, an adequate public health response demands comprehensive public health reporting and wider clinical recognition of long COVID (20). Candidate public health interventions to mitigate adverse sequelae of long COVID could include enhanced uptake by higher-risk groups of COVID-19 vaccines and non-pharmaceutical interventions including mask-wearing and improved ventilation to reduce the risk of acute infection (21,22), and expanded access to paid sick leave and disability insurance (16).

## Data Availability

All data produced in the present study are available upon reasonable request to the author.

## Notes

### Competing Interest Statement

The authors have declared no competing interest.

### Funding Statement

This study did not receive any funding.

### Author Declarations

All data are publicly available at: https://www.census.gov/programs-surveys/household-pulse-survey.html

### Summary of Updates

Analysis results have been updated for accuracy. New estimates also added on the economic burden of lost wages from long COVID.

